# Testing and Evaluation of Generative Large Language Models in Electronic Health Record Applications: A Systematic Review

**DOI:** 10.1101/2024.08.11.24311828

**Authors:** Xinsong Du, Zhengyang Zhou, Yifei Wang, Ya-Wen Chuang, Yiming Li, Richard Yang, Wenyu Zhang, Xinyi Wang, Xinyu Chen, Hao Guan, John Lian, Pengyu Hong, David W. Bates, Li Zhou

## Abstract

**Background:** The use of generative large language models (LLMs) with electronic health record (EHR) data is rapidly expanding to support clinical and research tasks. This systematic review synthesizes current strategies, challenges, and future directions for adapting and evaluating generative LLMs in EHR analyses and applications.

**Methods:** We followed the PRISMA guidelines to conduct a systematic review of articles from PubMed and Web of Science published between January 1, 2023, and November 9, 2024. Studies were included if they used generative LLMs to analyze real-world EHR data and reported quantitative performance evaluations. Through data extraction, we identified clinical specialties and tasks for each included article, and summarized evaluation methods.

**Results:** Of the 18,735 articles retrieved, 196 met our criteria. Most studies focused on Radiology (26.0%), Oncology (10.7%), and Emergency Medicine (6.6%). Regarding clinical tasks clinical decision support has the most studies of 62.2%, while summarizations and patient communications have the least studies of 5.6% and 5.1% separately. In addition, GPT-4 and ChatGPT were mostly used generative LLMs, which were used in 60.2% and 57.7% of studies, respectively. We identified 22 unique non-NLP metrics and 35 unique NLP metrics. Although NLP metrics have better scalability, none of the metrics were identified as having a strong correlation with gold-standard human evaluations.

**Conclusion:** Our findings highlight the need to evaluate generative LLMs on EHR data across a broader range of clinical specialties and tasks, as well as the urgent need for standardized, scalable, and clinically meaningful evaluation frameworks.

## 1. Introduction

Transformer architecture, introduced by Vaswani et al. in 2017, revolutionized artificial intelligence (AI) and natural language processing (NLP) by enabling efficient parallel processing of input sequences through self-attention mechanisms. This design allows models to capture long-range dependencies and contextual relationships more effectively than prior approaches.^1^ Large language models (LLMs) built on the Transformer architecture vary in design depending on their intended applications. Models like BERT,^2^ Longformer,^3^ NYUTron,^4^ GatorTron^5^ use only the encoder component of the Transformer architecture to model bidirectional context and generate dense representations that capture semantic and linguistic nuances. They perform well for language understanding tasks such as classification and named entity recognition. In contrast, generative models are designed to produce coherent text and are typically categorized as decode-only and encoder-decoder models. Decoder-only models, such as the GPT and Llama series,^2^ rely on the Transformer’s decoder architecture and are trained using autoregressive (next-token prediction) objectives. They are well-suited for tasks such as content generation, dialogue systems, and reasoning. Encoder-decoder models, such as T5 and BART, combine both encode and decoder components to transform input sequences into target outputs, supporting tasks like machine translation and summarization.

With the release of ChatGPT^6^ in November 2022, transformer-based generative LLMs have significantly accelerated progress in NLP and AI.^1,2,7^ These models, characterized by large parameter counts and autoregressive architectures, exhibit remarkable capabilities in language generation, reasoning, and contextual understanding, leading to widespread adoption in academia and industry.^8^ Despite increasing interest in healthcare, real-world deployment of generative LLMs for electronic health record (EHR) analyses and applications remains limited.^9^ Yet, these models offer unique potential to synthesize, summarize, and interpret unstructured clinical text, with applications in clinical decision support, documentation automation, and patient-provider communication.

However, translating generative LLMs into clinical practice requires rigorous testing and evaluation. Despite rapid progress, there remains a lack of systematic understanding of how these models are applied to real-world EHR data and how their performance is assessed. To address this gap, we conducted a systematic review to examine current applications, evaluation strategies, and remaining challenges. By synthesizing existing evidence, we aim to identify strengths, limitations, and opportunities that can inform the responsible and effective deployment of generative LLMs in clinical care.

## 2. Methods

### 2.1. Data Sources and Searches

We followed the Preferred Reporting Items for Systematic Review and Meta-Analyses (PRISMA) guidelines and conducted systematic searches in PubMed and Web of Science (**Figure 1)**.^10^ The process involved several key steps: a Boolean search, removal of duplicates, screening of studies, and data extraction. The Boolean search was conducted on November 09, 2024, using search terms and filters developed through team discussions. The search included LLM-related terms (e.g., “large language model”, “prompt engineering”) and the names of specific LLMs. The complete search query is provided in **Table 1**.

**Figure 1.**
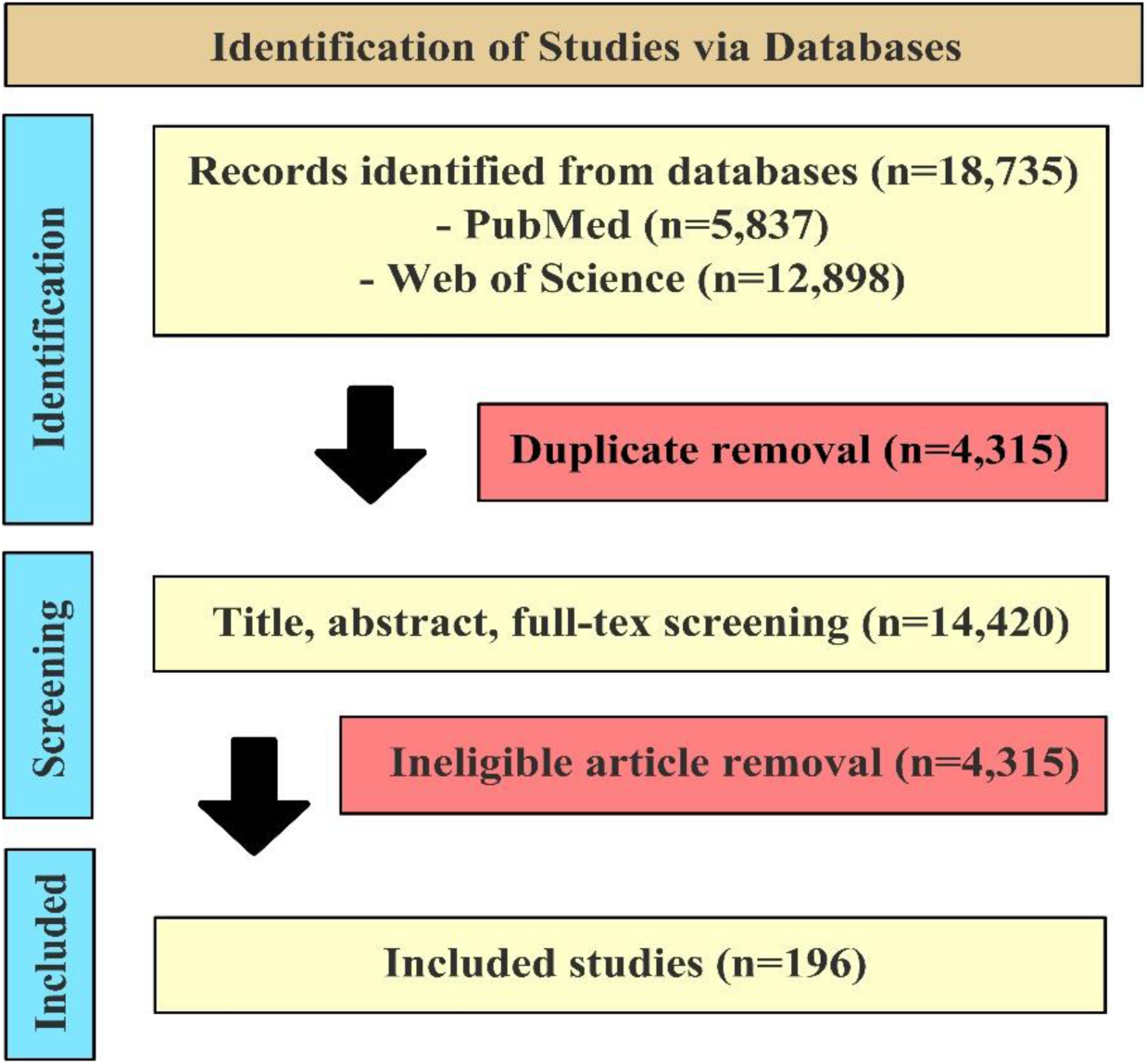
**PRISMA Flow Chart for Eligibility Screening**. A total of 18,735 records were identified from two databases: PubMed (n=5,837) and Web of Science (n=12,898). After removing 4,315 duplicates, 14,420 records underwent title, abstract, and full-text screening. Among them, 10,104 studies were excluded for not applying generative LLMs to real-world EHR data. Finally, 196 studies were included.

**Table 1.**
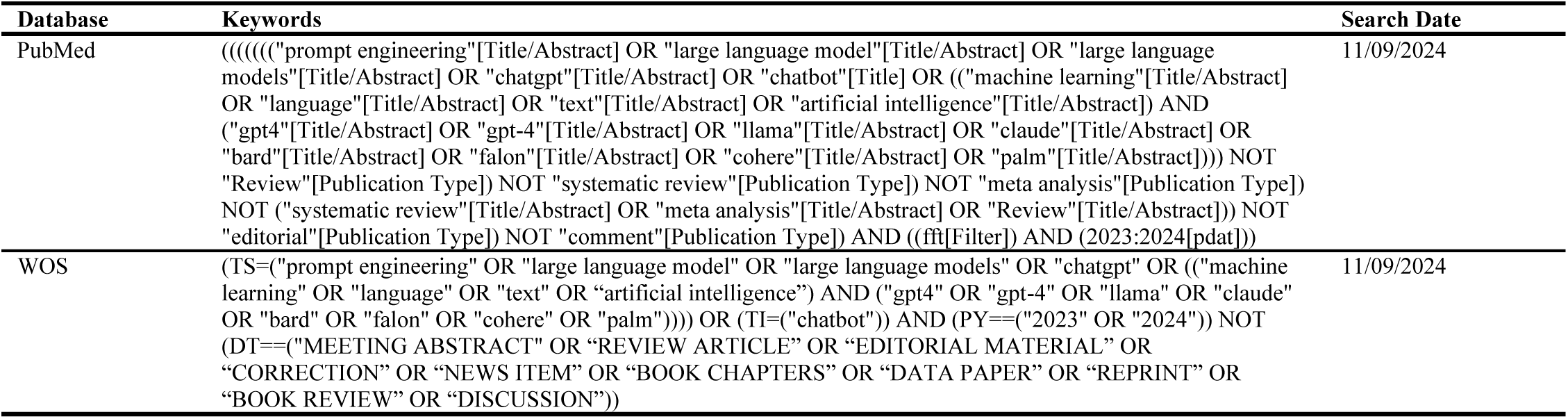
Search Keywords and Databases.

| Table 1. Search Keywords and Databases |  |  |
| --- | --- | --- |
| Database | Keywords | Search Date |
| PubMed | (((((("prompt engineering"[Title/Abstract] OR "large language model"[Title/Abstract] OR "large language models"[Title/Abstract] OR "chatgpt"[Title/Abstract] OR "chatbot"[Title] OR ("machine learning"[Title/Abstract] OR "language"[Title/Abstract] OR "text"[Title/Abstract] OR "artificial intelligence"[Title/Abstract]) AND ("gpt4"[Title/Abstract] OR "gpt-4"[Title/Abstract] OR "llama"[Title/Abstract] OR "claude"[Title/Abstract] OR "bard"[Title/Abstract] OR "falcon"[Title/Abstract] OR "cohere"[Title/Abstract] OR "palm"[Title/Abstract]))) NOT "Review"[Publication Type]) NOT "systematic review"[Publication Type]) NOT "meta analysis"[Publication Type]) NOT ("systematic review"[Title/Abstract] OR "meta analysis"[Title/Abstract] OR "Review"[Title/Abstract])) NOT "editorial"[Publication Type]) NOT "comment"[Publication Type]) AND ((ft[Filter]) AND (2023:2024[pdat])) | 11/09/2024 |
| WOS | (TS=("prompt engineering" OR "large language model" OR "large language models" OR "chatgpt" OR ("machine learning" OR "language" OR "text" OR "artificial intelligence") AND ("gpt4" OR "gpt-4" OR "llama" OR "claude" OR "bard" OR "falcon" OR "cohere" OR "palm")))) OR (TI=("chatbot")) AND (PY==("2023" OR "2024")) NOT (DT==("MEETING ABSTRACT" OR "REVIEW ARTICLE" OR "EDITORIAL MATERIAL" OR "CORRECTION" OR "NEWS ITEM" OR "BOOK CHAPTERS" OR "DATA PAPER" OR "REPRINT" OR "BOOK REVIEW" OR "DISCUSSION")) | 11/09/2024 |

### 2.2. Literature Screening

To focus on research articles presenting original data and quantitative results, we excluded certain article types, such as reviews. Given that the initial release of ChatGPT was on November 30^th^, 2022, we included articles published from 2023 onward, with only peer-reviewed articles included, while preprints were excluded.

The selection process began with the removal of duplicate articles, followed by a manual review of the deduplicated list. We excluded articles that met one or more of the following criteria: (1) Articles that were not of the appropriate type (e.g., preprints, reviews, editorials, comments). (2) Articles that did not involve generative LLMs; for example, those discussing chatbots but did not utilize LLMs were not considered. Although some encoder-based models like Longformer^3^, NYUTron^4^, GatorTron^5^ are powerful and widely used, studies involving only these encoder-based models were excluded. (3) Articles where the LLM was not used for English-language communication. (4) Articles where the LLM application was unrelated to healthcare (e.g., LLMs used for passing exams or conducting research). (5) Articles that lacked quantitative evaluation (e.g., those that only presented communication records with ChatGPT). (6) Articles that did not involve EHR data. (7) Articles where the EHR data used were not original (e.g., synthetic or summarized data).

During the eligibility screening process, two reviewers initially screened a set of 50 identical articles. If the agreement rate was above 90%, the reviewers proceeded to independently screen the remaining articles. If not, they discussed and screened an additional 50 articles until the agreement reached 90%.

### 2.3. Data Extraction

For included studies, we extracted their clinical tasks, clinical specialty, used large language models, and evaluation methods. We used heatmaps and bar charts to highlight the frequency of studies in each clinical task and specialty, as well as the frequency of LLM usage. We also extracted and summarized unique evaluation methods, provided use cases for each evaluation method, and summarized evaluation performances.

## 3. Results

Out of 18,735 unique records, 196 studies applied generative LLMs to EHR since 2023 (**Figure 1**). Each study was categorized into one clinical specialty. In addition, we categorized included articles into one or more of the following clinical tasks: 1) Clinical Decision Support (e.g., generating suggestions for diagnosis or treatment based on a patient’s history); 2) Information Extraction (e.g., extracting clinical entities and relationships from free-text clinical notes); 3) Documentation (e.g., generating clinical reports, notes or referral letters) 4) Summarization (e.g., summarizing longitudinal patient records); 5) Patient Communications (e.g., developing a conversational agent that answers clinical queries from patients). We also extracted generative LLMs used in the study.

### 3.1. Distribution of Studies Based on Clinical Specialty and Clinical Tasks

As illustrated in **Figure 2**, Radiology has the highest popularity with 51 (26.0%) articles, followed by Oncology (10.7%) and Emergency Medicine (9.7%). In terms of clinical tasks, clinical decision support has the highest frequency of 122 (62.2%), while summarization and patient communication have only 11 (5.6%) and 10 (5.1%) studies. In terms of the intersection of clinical specialty and clinical tasks, we found performing documentation tasks in Radiology has the most articles of 28 (14.3%), followed by clinical decision support in Emergency Medicine and clinical decision support in Radiology, which have 14 (7.1%) and 13 (6.6%) articles.

**Figure 2.**
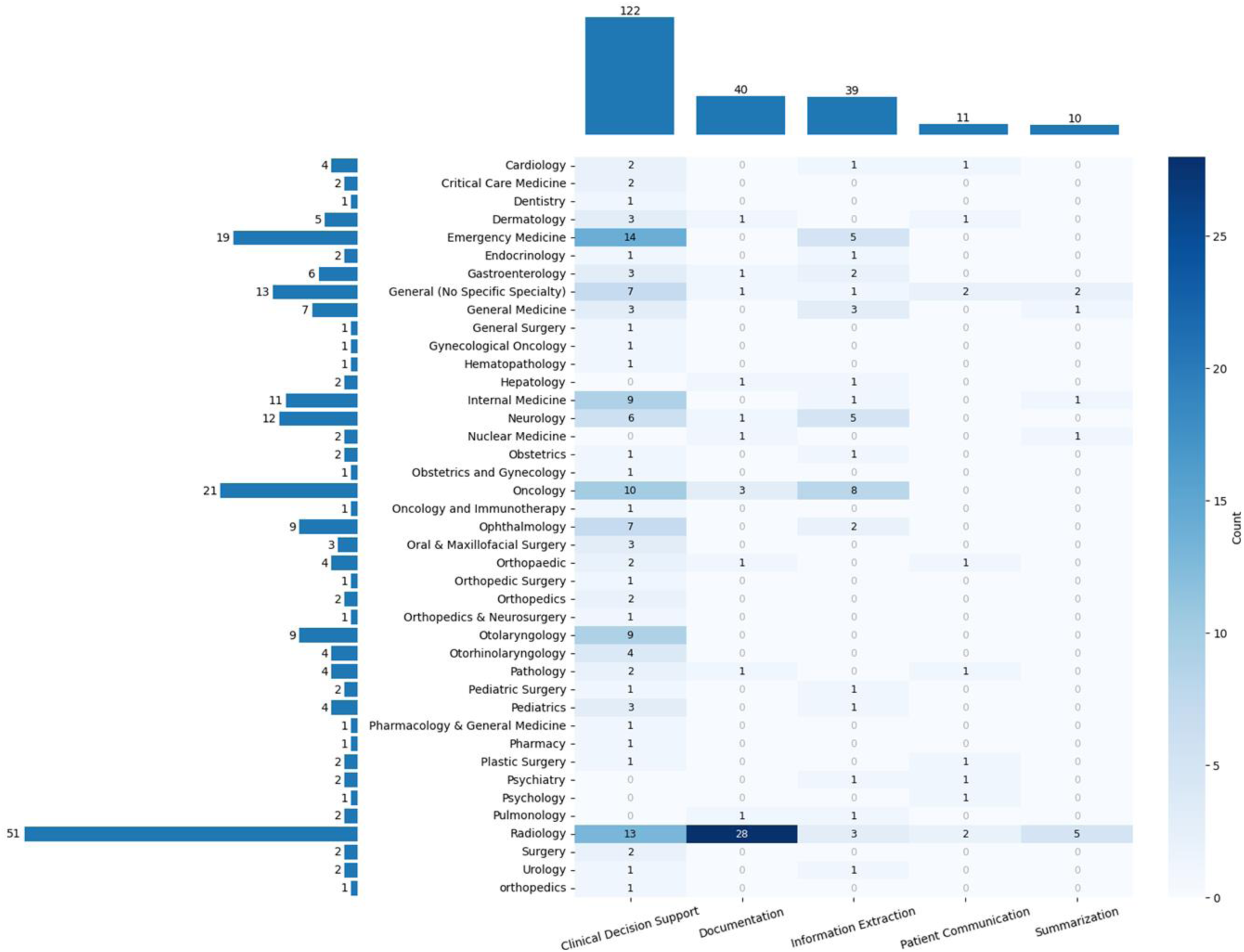
Distribution of Clinical Specialties and Tasks in Included Studies. The horizontal bar chart on the left illustrates the frequency of clinical specialties, with Radiology accounting for the largest share (n = 51, 26.0%). The top bar chart displays the distribution of clinical tasks, showing that the majority of studies (n = 122, 62.2%) focused on clinical decision support. The heatmap visualizes the intersection of clinical specialties and tasks, with the highest concentration of studies (n = 28, 14.3%) applying generative LLMs to documentation tasks in Radiology.

### 3.2. Usage of Generative LLMs

Figure 3 represents the LLM usage in different clinical specialties and tasks. GPT-4 has the highest use frequency of 114 (58.1%), followed by ChatGPT-3.5 (n=110, 56.1%), Llama-2 (n=18, 8.2%), and Gemini (n=16, 8.2%). Regarding clinical specialties, using GPT-4 in Radiology has the greatest number of studies of 30 (15.3%), followed by using ChatGPT in Radiology (n=23, 11.7%) and using ChatGPT in Oncology (n=14, 7.1%). In terms of clinical tasks, using ChatGPT for clinical decision support has the highest number of studies of 65 (33.2%), followed by using GPT-4 for clinical decision support (n=59, 30.1%) and using GPT-4 for documentation (n=26, 13.3%).

**Figure 3.**
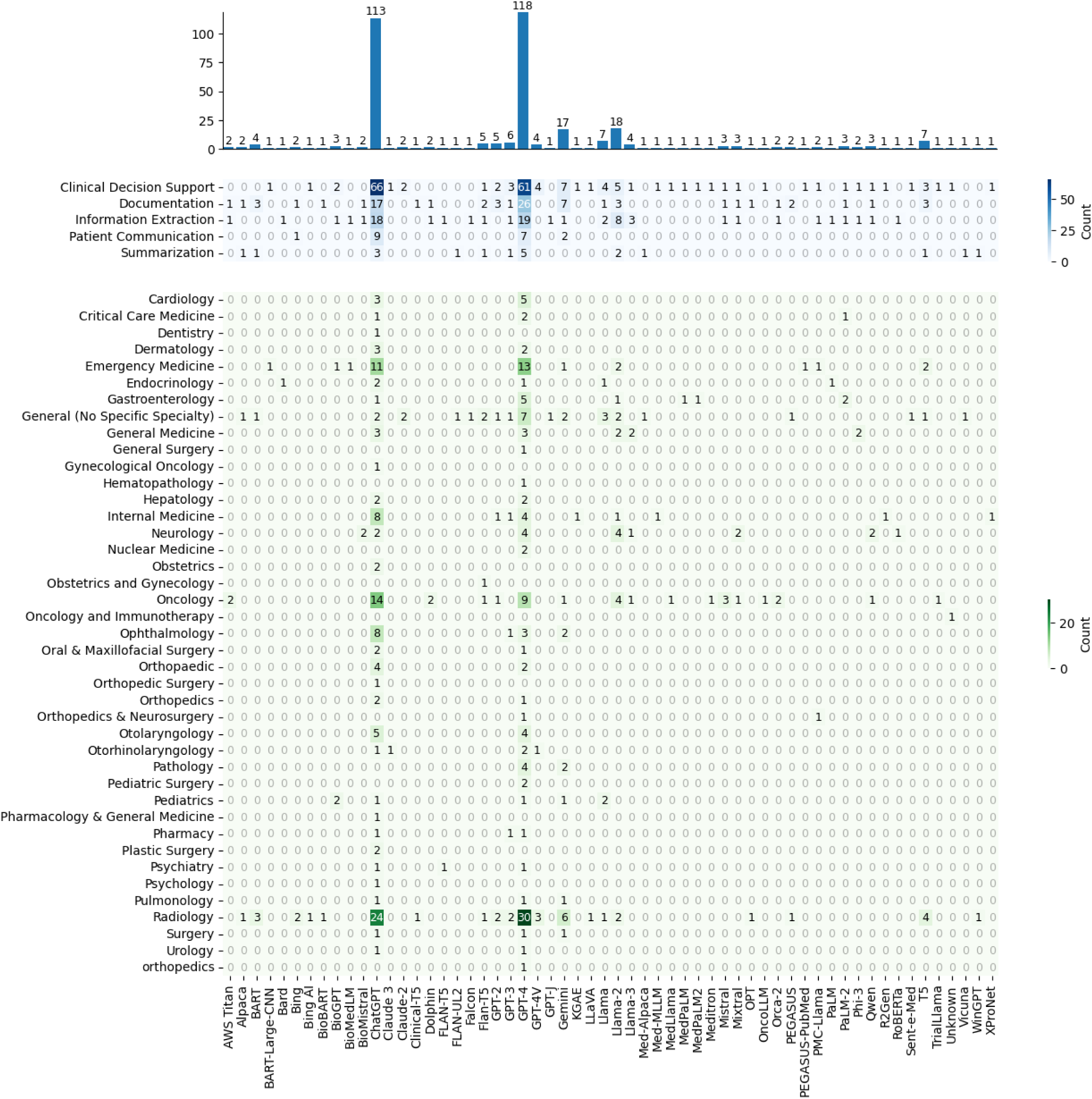
Usage of Generative LLMs Across Clinical Tasks and Specialties. The top bar chart displays the frequency with which each generative LLM was used in the included studies, with GPT-4 (n = 118, 60.2%) and ChatGPT (n = 113, 57.7%) being the most frequently used models. The upper heatmap shows the distribution of LLMs across different clinical tasks, revealing that ChatGPT was most commonly used for clinical decision support (n = 66, 33.7%), followed by GPT-4 for clinical decision support (n = 61, 31.1%) and GPT-4 for documentation tasks (n = 26, 13.3%). The lower heatmap presents the use of generative LLMs across clinical specialties, with GPT-4 in Radiology appearing most frequently (n = 30, 15.3%), followed by ChatGPT in Radiology (n = 24, 12.2%) and ChatGPT in Oncology (n = 14, 7.1%).

### 3.3. Evaluation Techniques

**Table 2** provides a summary of all unique evaluation metrics (non-NLP metrics) used in the included studies. A total of 22 different evaluation metrics were identified. In addition, we also found that 35 unique NLP metrics were used for evaluation, and three studies calculated their correlation with traditional evaluation methods, as shown in **Table 3**.^11–13^ However, none of the NLP metrics correlate with gold standard human evaluation.

**Table 2.**
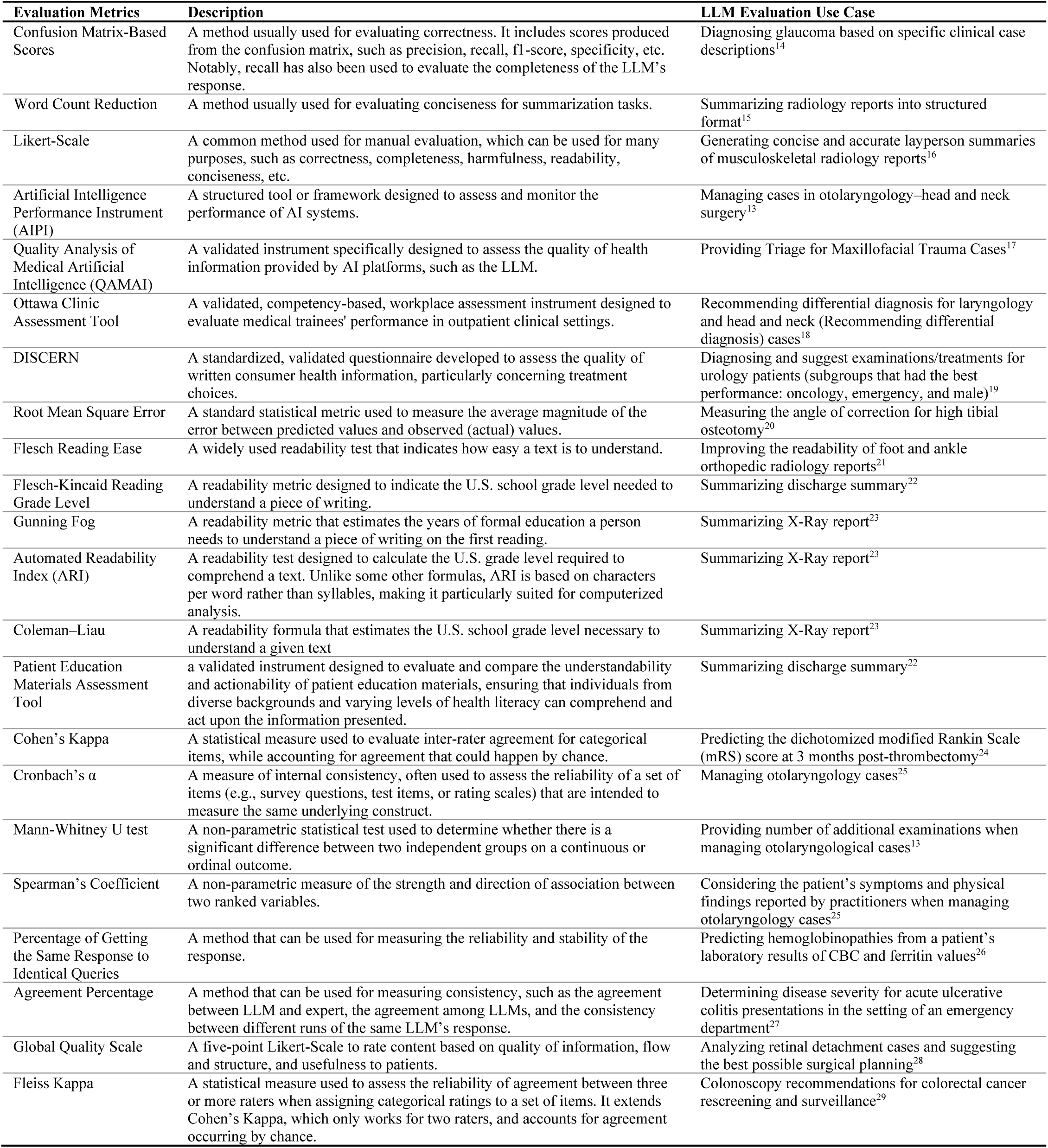
Glossary of Metrics for Generative LLM Manual Evaluation.

**Table 3.** Automated Metrics Used in Included Studies for Generative LLM Evaluation and Their Correlation (If Reported) with Human Evaluation.

| Evaluation Matrices | Correlated Evaluation (If Reported) Measured by Spearman’s Coefficient |  |
| --- | --- | --- |
|  | Reported Coefficient | Evaluation and Task |
| BLEU | 0.412 | Quality score of Impression generation for whole-body PET reports<sup>11</sup> |
|  | 0.225 | Completeness of clinical text summarization<sup>12</sup> |
|  | 0.125 | Correctness of clinical text summarization<sup>12</sup> |
|  | 0.075 | Conciseness of clinical text summarization<sup>12</sup> |
| BLEU-2 |  |  |
| BLEU-3 |  |  |
| BLEU-4 |  |  |
| ROUGE-1 | 0.402 | Quality score of Impression generation for whole-body PET reports<sup>11</sup> |
| ROUGE-2 | 0.379 | Quality score of Impression generation for whole-body PET reports<sup>11</sup> |
| ROUGE-L | 0.22 | Completeness of clinical text summarization<sup>12</sup> |
|  | 0.16 | Correctness of clinical text summarization<sup>12</sup> |
|  | 0.19 | Conciseness of clinical text summarization<sup>12</sup> |
|  | 0.398 | Quality score of Impression generation for whole-body PET reports<sup>11</sup> |
| BERTScore-Precision |  |  |
| BERTScore-Recall |  |  |
| BERTScore-F1 | 0.18 | Completeness of clinical text summarization<sup>12</sup> |
|  | 0.18 | Correctness of clinical text summarization<sup>12</sup> |
|  | 0.24 | Conciseness of clinical text summarization<sup>12</sup> |
|  | 0.407 | Quality score of Impression generation for whole-body PET reports<sup>11</sup> |
| MEDCON | 0.125 | Completeness of clinical text summarization<sup>12</sup> |
|  | 0.175 | Correctness of clinical text summarization<sup>12</sup> |
|  | 0.15 | Conciseness of clinical text summarization<sup>12</sup> |
| CIDEr | 0.194 | Quality score of Impression generation for whole-body PET reports<sup>11</sup> |
| BARTScore+PET | 0.568 |  |
| PEGASUSScore+PET | 0.563 |  |
| T5Score+PET | 0.542 |  |
| UniEval | 0.501 |  |
| BARTScore | 0.474 |  |
| CHRF | 0.433 |  |
| Moverscore | 0.420 |  |
| ROUGE-WE-1 | 0.403 |  |
| ROUGE-LSUM | 0.397 |  |
| ROUGE-WE-2 | 0.396 |  |
| METEOR | 0.388 |  |
| ROUGE-WE-3 | 0.385 |  |
| RedGraph | 0.384 |  |
| PRISM | 0.369 |  |
| ROUGE-3 | 0.345 |  |
| S<sup>3</sup>-pyr | 0.302 |  |
| S<sup>3</sup>-resp | 0.301 |  |
| Stats-novel trigram | 0.292 |  |
| Stats-density | 0.280 |  |
| BLANC | 0.165 |  |
| Stats-compression | 0.145 |  |
| SUPERT | 0.082 |  |
| Stats-coverage | 0.078 |  |
| SummaQA | 0.075 |  |

## 4. Discussion

Generative LLMs have shown exceptional potential across NLP tasks, yet their deployment on real-world EHR remains at an early stage. In this review, we examined LLM’s applications on real-world EHR data across diverse clinical tasks and summarized their evaluation techniques and performances. Our quantitative overview shows that using generative LLMs and real-world EHR has mostly focused on Radiology (26.0%), Oncology (10.7%), and Emergency Medicine (6.6%), but remains underexplored in other clinical specialties. Additionally, we found that summarization (5.6%) and patient communication (5.1%) are rarely investigated, highlighting the need for more future research. Moreover, most studies used GPT-4 (58.2%) and ChatGPT (56.1%), other generative LLMs should be tested and evaluated more frequently in the future as more and more LLMs are coming out recently.

Our review indicates a pressing need for standardized evaluation metrics and solutions to reduce the labor-intensive nature of human evaluation. We found that different studies often use varying metrics to achieve the same evaluation goals, highlighting the necessity of establishing standardized metrics for each evaluation purpose (e.g., correctness, completeness, etc.) to benchmark performance consistently. Although expert evaluation is considered the gold standard, it is impractical for physicians to thoroughly review all LLM outputs for performance evaluation.^11^ As data sizes increase, manual review becomes increasingly labor-intensive, costly, and time-consuming. Some studies started calculating correlations between automated similarity metrics and human subjective evaluation metrics, but none of the NLP metrics were found to correlate with human evaluation.

This review has several strengths, including its timely focus on the post-ChatGPT era, its exclusive inclusion of real-world EHR-based studies, and its comprehensive coverage of evaluation methods. By analyzing peer-reviewed articles across diverse clinical tasks and data modalities, our work offers a current and practical synthesis for researchers and implementers seeking to test and evaluate generative LLMs in healthcare applications. However, several limitations should be acknowledged. The studies included are drawn from a rapidly evolving and relatively new field, meaning that some findings are preliminary and potentially subject to publication bias, favoring positive results. The heterogeneity of clinical tasks, models, and evaluation metrics also limited our ability to conduct quantitative meta-analysis, and the focus on peer-reviewed literature from 2023 onward may have excluded valuable insights from preprints or earlier work. Looking ahead, future research should prioritize testing and evaluation of generative LLMs in a broader range of clinical specialties and tasks, and standardize evaluation methods. Collectively, these directions will be vital to support the responsible and effective adoption of generative LLMs in clinical settings.

## 5. Conclusion

This systematic review highlights the rapid but uneven growth in applying generative LLMs to real-world EHR data. Current research is concentrated in a few clinical specialties—most notably Radiology—and heavily focused on decision support tasks, while critical areas such as summarization and patient communication remain underexplored. GPT-4 and ChatGPT are the most frequently used models in existing studies, underscoring the need to broaden evaluations to include a wider range of generative LLMs. Standardized and clinically meaningful evaluation frameworks are lacking, and current NLP metrics show limited correlation with expert assessment. To advance the responsible integration of generative LLMs into healthcare, future work should expand evaluations across diverse clinical settings and tasks, and prioritize the development of scalable, reliable evaluation methodologies.

## Data Availability

All data produced in the present work are contained in the manuscript

## Acknowledgments

This study was funded by NIH-NLM 1R01LM014239, NIH-NIA R01AG080429, and OpenAI Researcher Access Program.

## Author Contributions

Conceptualization and design: XD, ZZ, YW, LZ. Deciding search databases and keywords: XD, LZ. Literature search: XD. Literature screening: XD, ZZ, YW, YL, RY, WZ, XW, XC, HG, JL. Data extraction: XD, ZZ, YW, YWC, YL. Original draft preparation: XD. Visualization: XD, ZZ, YW. Critical feedback for revision: ZZ, YW, DWB, ZZ. Funding acquisition: XD, LZ. Supervision: LZ.

